# Adherence to Pre-exposure Prophylaxis intervention by transgender women: A systematic review

**DOI:** 10.1101/2022.12.20.22283755

**Authors:** Jorge Eduardo Moncayo-Quevedo, María Del Mar Pérez-Arizabaleta, Lina María Villegas-Trujillo, Alejandra Rodríguez-Ortiz

## Abstract

**Background:** The prevalence of HIV is higher in the transgender population. Recently, the preexposure prophylaxis (PrEP) intervention has been proven successful in reducing HIV acquisition in trials among men who have sex with men (MSM), and heterosexual couples. This research aims to investigate the adherence to PrEP by HIV-negative transgender women (TW).

**Methods:** Were followed the Cochrane Handbook for Systematic Reviews of Interventions and the PRISMA Statement. Research in WoS, Ovid, Scopus, MEDLINE, and the Cochrane Central Register of Controlled Trials (CENTRAL) electronic databases for studies that involved HIV-negative TW population and focused on their adherence to PrEP intervention and condom use after the treatment.

**Results:** 11 studies were included. TW sample sizes were low in comparison to the total sample, which often included men who have sex with men (MSM) population. The participation and adherence to the intervention was low compared to MSM, and it was measured mainly by self-report (72.7%) or by Tenofovir-diphosphate (TFV-DP)/ Emtricitabine triphosphate (FTC-TP) dried blood spot (DBS) (45.5%).

**Conclusions:** It is important to increase awareness and explain the effect of PrEP on feminizing hormone therapy at the beginning of the trials. Nevertheless, the low adherence may be affected by the interaction between drugs and the barriers faced to use the health services.

## Introduction

Transgender populations refer to individuals whose gender identity or expression differs from that associated with their sex at birth; hence, ‘transgender women’ can describe natal males who have a feminine gender identity or expression.^1^ The prevalence of human immunodeficiency virus (HIV) is higher in the transgender population than among the general population,^2^ even among men who have sex with men (MSM), as seen in a meta-analysis made in the United States which reports a HIV prevalence rate among transgender women up to 27.7%.^3^ Operario et al.^4^ found in their systematic review that overall crude HIV prevalence among transgender women sex workers was 27.3%, meanwhile among transgender women not engaged in sex work was 14.7%. It has also been reported that transgender women who engage in sex work are at high risk for HIV compared to natal male and female sex workers. According to Poteat et al.^5^ by 2015, there were no evidence-based prevention interventions focused exclusively on transgender women. More recently, the preexposure prophylaxis (PrEP) intervention has been proven successful in reducing HIV acquisition in trials among MSM,^6^ and heterosexual couples,^7^ among others. PrEP consists of a daily drug regimen of tenofovir disoproxil fumurate and emtricitabine. However, there are concerns that the HIV prevention that benefits the population that uses PrEP might decline the widespread condom use across these populations at risk of HIV.^8,9^ An additional concern is a possible reduction in the use of PrEP due to a representation of reduced effectiveness of feminizing drugs when combined. In this context, in this systematic review, we aim to investigate about the adherence to PrEP by HIV-negative transgender women.

## Methods

This systematic review followed the recommendations made by the Cochrane Handbook for Systematic Reviews of Interventions,^10^ and the PRISMA Statement 2020.^11^ The protocol for this study was registered in OSF (https://osf.io/ukdyz/)

### Eligibility Criteria

We searched for randomized controlled trials and observational case-control studies that worked with the HIV-negative transgender women population and focused on their adherence to PrEP intervention, regardless of race or country. We selected articles that included transgender women in their samples either alone or in combination with MSM population as long as in the results section the data was sepatered for both. Only studies that had the purpose of reducing risky behavior and increasing safe behaviors were included. Studies that included only MSM population and transgender men were excluded.

### Information sources and search

We conducted a systematic search in the WoS, Ovid, Scopus, MEDLINE, and the Cochrane Central Register of Controlled Trials (CENTRAL) electronic databases, with no language restriction and no date restriction until March 2022. For the search of the literature, we used MeSh terms. We used a combination of MeSH terms and non-controlled vocabulary that we considered crucial to our objective in the equation:

((“HIV”[Title/Abstract] OR “human immunodeficiency virus” [Title/Abstract])

AND

“transgender women”[Title/Abstract]

AND

(“Pre-exposure prophylaxis”[Title/Abstract] OR “PrEP”[Title/Abstract]))

AND

(Clinical study [Filter] OR clinical trial [Filter] OR comparative study [Filter] OR observational study [Filter] OR randomized controlled trial [Filter])

We also conducted a generic and academic Internet search and a metasearch to control publication bias. A search strategy defined for “gray literature” was included to gather information from Google Scholar.

### Selection of studies

Eligibility assessment was performed independently in an unblinded standardized manner by two reviewers. Titles and abstracts of all citations were identified, and all the potentially eligible studies were selected. The same authors then independently evaluated the complete text versions of these articles to determine whether each study fulfilled the inclusion criteria. A two-reviewer dissolved conflict and disagreements were resolved by consensus, and where dissent could not be solved.

### Data extraction and synthesis of results

The abstraction variables included author, year, country, study design, sample size, participant demographics, an additional type of interventions (besides PrEP), length of follow-up, prophylaxis, sex workers or stable partner, and Odds Ratio (OR) of adherence to PrEP. A data extraction table was made in Microsoft Excel to organize the results.

Two authors independently extracted data from eligible studies. The primary outcome sought for this review was the adherence to PrEP by HIV-negative transgender women. Two researchers reviewed each study found in the databases by title and abstract, selecting the more adequate ones. Subsequently, they reviewed full-texts of previously selected articles and screened them according to the inclusion criteria. With the studies finally set, we extracted the data. Disagreements were resolved by consensus.

### Risk of bias assessment

Two authors independently assessed the methodological quality of data and risk of bias of the included studies using the Newcastle-Ottawa Scale (NOS), including nine items. The selection criteria contain four items: 1) the adequate case definition, 2) representativeness of the cases, 3) controls selection, 4) controls definition. The comparability criteria include comparability of cases and controls according to the design or analysis. The exposure criteria contain three aspects: first, ascertainment of exposure; second, the same method of ascertainment for cases and controls and third, non-response rate. Disagreements were solved through discussion until consensus. A study scoring six or higher was considered to be of sufficient quality.

### Strategy for data analysis

For dichotomous outcomes, the number of events and the number of participants in each group (intervention or control) were extracted. Odds ratios (OR) or hazard ratios (RR) (both raw and adjusted ratios, if available) were removed, along with their 95% confidence intervals (CI) and p-values. For continuous results, we extracted data from means, standard deviations, and the number of participants in each group. Data from medians, ranges, and p-values were extracted from nonparametric tests for continuous asymmetric data. We took out percentages, mean or median scores for change from baseline for controlled before and after studies. If the change scores were not available, we extracted the post-intervention values. Any discrepancy between the two investigators was resolved by discussion or consultation with other co-authors of the systematic review.

## Results

### Selection of studies

The literature search strategy yielded 700 potentially eligible articles. Subsequently, after discarding duplicate and review full-text articles, 11 fulfilled the inclusion criteria and were included in the systematic review (Figure 1).

**Figure 1.**
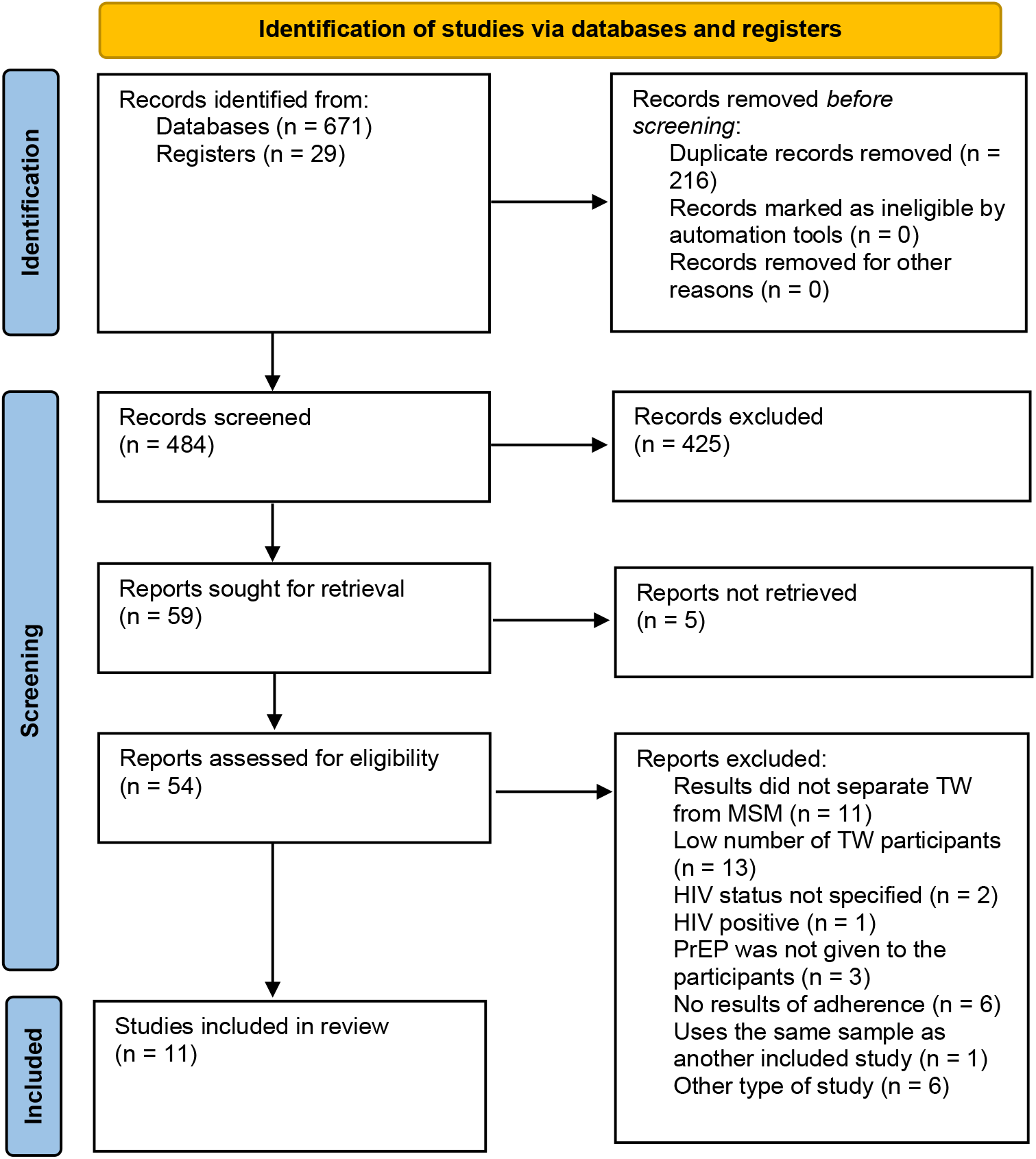
PRISMA flow diagram 2020.

### Characteristics of included studies

The predominant study design was prospective cohort studies (81.8%). Most included studies were from Thailand (36.4%), the United States (36.4%), and Brazil (27.3%) between 2014 and 2021 (Table 1). Transgender women’s sample sizes from the studies were small compared to the total sample size, which often included the MSM population; considering a total of 8261 participants, only 815 were transgender women, accounting for 9.9%. The age of the participants was 18 or older. However, we noticed that most studies reported the age of transgender women and MSM population combined. Only three studies reported separate age data for transgender women (27.3%), and the follow-up period went from 1 up to 18 months. We did not perform a meta-analysis due to the heterogeneity of the studies.

**Table 1.** Characteristics of studies included.

| Author | Year | Country | Sudy design | Full<br>sampl<br>e size | TW<br>Sampl<br>e size | Mean Age<br>(years) | Additional<br>intervention<br>s | Follow-up (months) |
| --- | --- | --- | --- | --- | --- | --- | --- | --- |
| Connolly<br>et al.<sup>12</sup> | 2020 | United States | Cohort study | 38 | 19 | 26* | No | 3 |
| Grant et al.<sup>13</sup> | 2014 | United States, Brazil, Peru,<br>Ecuador, South Africa | Retrospective<br>cohort study | 1225 | 58 | >18* | No | 3 |
| Green et al.<sup>14</sup> | 2021 | Vietnam | Cohort study | 1131 | 62 | - | No | 18 |
| Hoagland<br>et al.<sup>15</sup> | 2017 | Brazil | Cohort study | 450 | 25 | >18* | No | 1 |
| Seekaew et al.<sup>16</sup> | 2019 | Thailand | Cohort study | 653 | 89 | 28.9* | No | > 3 |
| Spinelli et al.<sup>17</sup> | 2019 | United States | Retrospective<br>cohort study | 364 | 45 | >25* | No | 12 |
| Leu et al.<sup>18</sup> | 2019 | South Africa, Thailand, Peru,<br>United States, and Puerto<br>Rico | Cohort study | 187 | 23 | 31.4* | No | 2 |
| Kimani et al.<sup>19</sup> | 2021 | Kenya | Cohort study | 53 | 11 | >18 | No | - |
| Phanuphak<br>et al.<sup>20</sup> | 2018 | Thailand | Cohort study | 1697 | 230 | 26.1 | No | 12 |
| Ramautarsing et al.<sup>21</sup> | 2020 | Thailand | Cohort study | 2088 | 232 | 26 | No | 3 |

\*The reported age in the study was for all key populations evaluated, not exclusively for the transgender women population. TW: Transgender women. PrEP: Preexposure prophylaxis.

### Outcome variables

Outcome variables are described in table 2. Most studies did not say if the participants were sex workers, three studies did, two of them from Thailand. The participants adherence to the intervention was low compared to the MSM population, and it was measured mainly in two forms: by self-report (72.7%) or by Tenofovir-diphosphate (TFV-DP)/ Emtricitabine triphosphate (FTC-TP) dried blood spot (DBS) (45.5%).

**Table 2.** Outcome variables from the included studies

| Author | Year | TW Sex worker n (%) | Measurement of adherence | OR/RR Adherence |
| --- | --- | --- | --- | --- |
| Connolly et al.<sup>12</sup> | 2020 | 14 (28%) | TFV-DP DBS assessment and self-reported PrEP adherence with a three-item validated measure. | RR 0.63 (0.20–1.95) P value 0.46 |
| Grant et al.<sup>13</sup> | 2014 | No | TFV-DP DBS assessment | OR 2.03 (1.27 to 3.25) P-value <0.0001 |
| Green et al.<sup>14</sup> | 2021 | - | Self-reported PrEP adherence | For TW, being aged >30 years was associated with continuing on PrEP (aOR 5.62 (95% CI: 1.05-29.9), P value 0.043). |
| Grinsztejn et al.<sup>22</sup> | 2018 | - | TFV-DP and FTC-TP DBS assessments | OR 0.83 (0.32–2.16) P-value 0.70 |
| Hoagland et al.<sup>15</sup> | 2017 | - | TFV-DP and FTC-TP DBS assessments using liquid chromatography and mass-spectroscopy (LC/MS/MS). | OR 0.75 (0.27-1.83) P-value 0.55 |
| Seekaew et al.<sup>16</sup> | 2019 | - | Self-reported PrEP adherence categorized into two levels: Low Adherence (no follow-up or $\leq 3$ pills/week) and Good Adherence ($\geq 4$ pills/week). | Low adherence associated with TW (aOR: 2.2, 95% CI: 1.27–3.83, p = 0.005) |
| Spinelli et al.<sup>17</sup> | 2019 | - | Self-reported PrEP adherence | Transgender women were more likely to discontinue PrEP early (aRR; 2.16; 95% confidence interval, 1.36–3.49). Late discontinuation: aRR 1.84 (.98–3.46) |
| Leu et al.<sup>18</sup> | 2019 | - | Self-reported PrEP adherence by short message service (SMS) | In the Daily oral regimen, MSM was more likely to be high-adherers than TW (OR=4.76, 95% CI=1.35, 16.67, p=0.015). |
| Kimani et al.<sup>19</sup> | 2021 |  | Self-reported PrEP adherence and TFV-DP DBS assessment | - |
| Phanuphak et al.<sup>20</sup> | 2018 | 76 (33%) | Self-reported PrEP adherence | OR 1 (Reference) P-value <0.001 |
| Ramautarsing et al.<sup>21</sup> | 2020 | 57 (24.6%) | Self-reported PrEP adherence | - |

TFV-DP: Tenofovir-diphosphate. FTC-TP: Emtricitabine triphosphate. DBS: Dried blood spot. NR: Not reported. TW: Transgender women. PrEP: Preexposure prophylaxis.

### Risk of bias assessment

Each study chosen for this systematic review was carefully evaluated according to the Newcastle-Ottawa scale, and the quality scores of the studies are shown in Table 3. All included studies showed moderate to high scores.

**Table 3.** Risk of bias assessment for included studies

| Author | Year | Selection | Comparability | Outcome | Total |
| --- | --- | --- | --- | --- | --- |
| Connolly et al.<sup>12</sup> | 2020 | 3 | 1 | 3 | 7 |
| Grant et al.<sup>13</sup> | 2014 | 4 | 1 | 3 | 8 |
| Green et al.<sup>14</sup> | 2021 | 3 | 1 | 3 | 7 |
| Grinsztejn et al.<sup>22</sup> | 2018 | 3 | 1 | 3 | 7 |
| Hoagland et al.<sup>15</sup> | 2017 | 3 | 1 | 3 | 7 |
| Seekaew et al.<sup>16</sup> | 2019 | 3 | 1 | 3 | 7 |
| Spinelli et al.<sup>17</sup> | 2019 | 3 | 1 | 3 | 7 |
| Leu et al.<sup>18</sup> | 2019 | 3 | 1 | 3 | 7 |
| Kimani et al.<sup>19</sup> | 2021 | 3 | 1 | 3 | 7 |
| Phanuphak et al.<sup>20</sup> | 2018 | 3 | 1 | 3 | 7 |
| Ramautarsing et al.<sup>21</sup> | 2020 | 3 | 1 | 3 | 7 |

## Discussion

This systematic review aimed to shed light on the adherence to PrEP by HIV-negative transgender women. Here we found lower participation in the studies^15^ and adherence to PrEP by transgender women than the MSM population. This could be explained by the concern that transgender women may have regarding the effect of PrEP on hormone therapy, as it was shown by Deutsch et al.^23^ who found lower TDF-DP concentrations in DBS among transgender women used feminizing hormones in comparison to the other participants. The authors explained this finding by lacking knowledge regarding the drug-drug interactions during the trial. Even though the drugs used in PrEP treatment are metabolized in the kidney, estrogens and progestogens are metabolized in the liver, no systemic drug-drug interactions are estimated,^24^ at the moment of the trial, there were no specific studies that proved these expectations.^23^ Marshall & Mimiaga^25^ emphasize the need for investigations to address any effects of PrEP interventions and the feminizing hormone therapy. They would provide additional assurance to transgender women participating in PrEP trials. Another reason behind the low participation rate in intervention studies is the constant discrimination they face from healthcare workers that have been previously reported.^26^ In this regard, education for healthcare providers on the LGBTQ+ community and their needs is key to gaining trust from transgender women,^27^ and creating a safe and comfortable environment to be more willing to uptake PrEP.^28^

Moreover, the PrEP awareness by transgender women is lower compared to the MSM population; according to the recent study by Wilson et al.^29^ the authors found that only 79.1% of transgender women knew about PrEP vs. 96.7% MSM. Those with previous knowledge believed that the treatment was only for MSM, seemingly focused on them.^30^ Likewise, Galka et al.^31^ found that only 20% of the Malaysian transgender women that partake in the investigation had previous knowledge of PrEP. Contrastingly, the study made by Horvath et al.^32^ in the United States found high awareness amongst transgender youth; however, only one participant was using PrEP.

From the results from this systematic review, we also noticed that it was more common than expected to see studies that did not differentiate the MSM population from the transgender women, clustering them as one population in the results section. Different authors have noticed this situation before, including Sevelius et al.^33^ and Wilson et al.^29^ and it is found in HIV research and the health care system.^34^ There are several reasons behind the need to differentiate both populations, including the unique barriers that transgender women encounter, such as lack of social support, anti-trans stigma, mental health issues, and gender-affirming hormones, among others that can affect negatively PrEP engagement.^35–39^ According to Sevelius et al.^34^ negative experiences with clinics and providers unsupportive of transgender identities may influence the decision of transgender women to participate in PrEP programs.

Sevelious et al.^33^ recently made one of the few studies that researched PrEP adherence in the transgender population exclusively and found that amongst this community, transgender women are more likely to adhere to the intervention during the follow-up. This is an interesting result, given that those differences cannot be found when combining the results from MSM with the transgender population. Another factor that differentiates the MSM population from transgender women is that amongst the latter, there is a high percentage of sex workers,^40^ where they face a higher rate of exposition to HIV infection than other key populations. This factor alone should be a reason to involve more transgender women in specifically designed PrEP research, not adapted from studies of MSM.^23^

### Conclusions

Our results showed a tendency of the studies to present results from transgender women and MSM populations altogether. However, considering that both populations have different backgrounds, we suggest that further studies show the results from both populations separated, allowing us to draw more accurate conclusions. Moreover, our findings regarding the low participation and adherence of transgender women in PrEP interventions suggest the importance of increasing awareness and give a clear explanation to this population concerning the effect of PrEP on feminizing hormone therapy at the beginning of the trials. However, further studies are needed to determine this potential effect. Furthermore, we suggest the inclusion of specific indicators of effectiveness, as the condom-use variable before and after the use of PrEP in future studies with transgender women.

## Supporting information

PRISMA checklist

## Data Availability

The authors confirm that the data supporting the findings of this study are available within the article and its supplementary materials.

## Authors’ Contributions

JEMQ, MDMPA, LMV, and ARO, contributed equally to the conception, design of the work, acquisition, analysis, interpretation of data, drafting the work, revising it critically for important intellectual content. All authors approved the final version of the manuscript and agreed to be accountable for all aspects of the work.

## Abbreviations used

CENTRAL: Cochrane Central Register of Controlled Trials
CI: Confidence Interval
DBS: Dried Blood Spot
FTC-TP: Emtricitabine triphosphate
HIV: Human Immunodeficiency Virus
MSM: Men who have Sex with Men
OR: Odds Ratio
PrEP: Preexposure Prophylaxis
RR: Risk Ratio
TFV-DP: Tenofovir-diphosphate

